# Clinical and Virologic Factors associated with Outcomes of COVID-19 before and after Vaccination among Veterans: Retrospective Analysis from Six New England States

**DOI:** 10.1101/2022.02.24.22271468

**Authors:** Megan Lee, Danielle Cosentino, Tassos C. Kyriakides, Tricia Cavallaro, Gary Stack, Shaili Gupta

**Affiliations:** Yale School of Medicine, New Haven, CT; Department of Clinical Informatics, VA Connecticut Healthcare System, West Haven, CT; Department of Veterans’ Affairs Cooperative Studies Program, VA Connecticut Healthcare System, West Haven, CT; Pathology and Laboratory Medicine Service, VA Connecticut Healthcare System, West Haven, CT; Department of Laboratory Medicine, Yale School of Medicine, New Haven, CT; Department of Internal Medicine, Yale School of Medicine, New Haven, CT; Department of Medicine, VA Connecticut Healthcare System, West Haven, CT

**Author notes:** **Corresponding Author:** Shaili Gupta, MBBS, Department of Medicine, Yale School of Medicine; VA Connecticut Healthcare System 950 Campbell Ave, Bldg 1, 5^th^ floor, Mailstop 111; West Haven, CT 06516.

## Abstract

**Background:** A region-wide analysis of COVID-19 outcomes in New England has not been done. We aimed to characterize clinical, demographic, and vaccination status affecting COVID-19 clinical outcomes and describe viral epidemiology.

**Methods:** Clinical variables of Veterans with COVID-19 in Veterans Administration healthcare systems in six New England states from April 8, 2020, to September 2, 2021 were correlated with outcomes of 30-day mortality, non-psychiatric hospitalization, intensive care unit admission (ICU-care), and post-vaccination infection. We sequenced 754 whole viral genomes and 197 partial genomes.

**Results:** Of 4,170 Veterans with COVID-19, 81% were White, 8% women, mean age was 60.1 ±17.7 years, and 2,399 became fully vaccinated. Overall, 19% Veterans needed hospitalization, 2.8% required ICU-care, and 3.7% died. Veterans with post-vaccination COVID-19 were older, with higher rates of tobacco/drug use, CKD, and malignancy, and 0.38% died. Among the unvaccinated, ICU-care and mortality correlated with age, while hospitalization correlated with age, male sex, black race, drug use, chronic heart disease, COPD, CKD, and chronic liver disease. Age, CKD, and alcohol use correlated with hospitalization in vaccinated patients.

Most New England Veterans (>97%) were infected with B.1 sub-lineages with the D614G mutation in 2020 and early 2021. B.1.617.2 lineage (71%) predominated after July 2021, including the post-vaccination infections.

**Conclusion:** In New England Veterans with mean age of 60 years, age and CKD significantly correlated with hospitalization regardless of vaccination-status. Age correlated with mortality and ICU-care among the unvaccinated. The Delta variant of SARS-CoV-2 (B.1.617.2) dominated post-vaccination infections.

## Introduction

The Coronavirus Disease 2019 (COVID-19) caused by Severe Acute Respiratory Syndrome Coronavirus-2 (SARS-CoV-2) has infected >40 million people in the United States and caused over 650,000 deaths.^1^ While some host and viral factors have been found to be associated with transmission rates, disease severity, and mortality, there are significant variations in these outcomes among specific populations.^2,3^ Some host-factors that have been shown to have a significant association with clinical outcomes include age, Charlson comorbidity index score, and body mass index (BMI).^4-9^ Viral factors have similarly been investigated for affecting outcomes. U.S. Center for Disease Prevention (CDC) and World Health Organization (WHO) have been diligently maintaining active surveillance of the evolving SARS-CoV-2 virus across the globe, and several mutations have been known to result in the generation of Variants of concern (VOC) and Variants of interest (VOI).^10,11^

Early during the pandemic course in the United States, viral isolates on the East Coast of the US were predominantly of the B.1 lineage that contained the Spike protein D614G substitution.^12,13^ This substitution has been suggested to have enhanced transmissibility,^14^ said to be supported by the higher upper airway viral loads and higher viral replication in lung epithelial cells.^15^ With the increasing focus on emerging SARS-CoV-2 genetic variants, and concerns about suboptimal protection from vaccines against infection by certain variants, more research is required on how mutations affect risk of infection, as well as Veteran outcomes.^16-20^ Both regional epidemiology of the virus, and unique clinical characteristics of certain populations, can play a role in determining outcomes of the infection.

The work presented here evaluated a unique regional population for factors associated with clinical outcomes of COVID-19. U.S. Veterans make a reasonably vulnerable population because of their average age of 58 years, which is higher than the average age of a U.S. resident (38.5 years), and because Veterans are largely men (89%).^21^ Several studies have linked male sex and higher age to poorer outcomes of COVID-19.^2,22-24^ In the beginning of the COVID-19 era, we conducted a small pilot study on Veterans with COVID-19 between April, 2020 and September, 2020 from the same region of New England, and found age, dementia and low BMI to be associated with mortality.^25^ Here we present the analysis of outcomes throughout the COVID-19 era in these Veterans from the New England states.

In a region-wide analysis of Veterans in New England, we aimed to (1) characterize clinical, demographic, and viral factors affecting Veteran outcomes, (2) conduct genomic evaluations of lineages and spike mutations throughout the COVID-19 era to determine evolution of SARS-CoV-2, and (3) understand clinical and genomic factors that impact morbidity and mortality risk in post-vaccination cases.

## Methods

We conducted a retrospective analysis of demographic and clinical variables to identify their relationship with outcomes of COVID-19 among Veterans in New England. The Veterans Affairs Connecticut Healthcare System (VACHS) in West Haven, Connecticut has carried out testing for SARS-CoV-2 on nasopharyngeal specimens from all six VA healthcare centers in New England states (Connecticut, Massachusetts, Maine, New Hampshire, Rhode Island, Vermont). The virus isolated from several samples testing positive was sent for whole genome sequencing (WGS), and genomic data so generated provided insight into the evolution of the virus in this large region. The VACHS Institutional Review Board (IRB) approved the creation and maintenance of a data repository of all Veterans in New England diagnosed with COVID-19 and a viral repository of the SARS-CoV-2 RNA received from all six New England facilities.

### Cohort

Our cohort included all Veterans receiving care at VA healthcare centers in six New England states from April 8, 2020, to September 2, 2021, who had a diagnosis of COVID-19 based on one of three four PCR-based tests: Xpert® Xpress SARS-CoV-2 (GeneXpert system; Cepheid), Simplexa® COVID-19 Direct Kit (LIAISON MDX; DiaSorin Molecular), BioFire® Respiratory 2.1 Panel (BioFire FilmArray; BioFire Diagnostics), and Roche ccobas® SARS-CoV-2 test (cobas 6800 system; Roche Diagnostics), and had accessible chart records.

### Data Collection

Data collection methods included utilizing Structured Query Language (SQL) to query the VA Corporate Data Warehouse (CDW) for SARS-CoV-2 completed lab test results to identify a Veteran cohort of confirmed SARS-CoV-2 cases. The specimens were all collected, performed and resulted at a VA New England facility utilizing the electronic laboratory package that is interfaced with the VA Computerized Patient Record System. Tests performed at VA facilities outside of this area or other institutions were not included in the cohort. The dataset included demographic variables [age, sex, race, body-mass index (BMI), and state of residence when diagnosed with COVID-19] as recorded in Veteran medical records. Health information was extracted for this cohort from the outpatient and inpatient datasets available. A unique identifier for each Veteran allowed for the datasets to be joined and organized by each distinct Veteran. This removed duplication of test results, health factors, admissions or pertinent information across data sources. ICD-9 and ICD-10 diagnostic codes for the following morbidities documented in the medical record were included: dementia, alcohol use, tobacco use, drug use (cannabis, opioid, cocaine, hallucinogen, amphetamine, or anxiolytic), chronic heart disease [including coronary artery disease (CAD), cardiomyopathy, atrial fibrillation, other arrhythmia, congestive heart failure, and cerebrovascular accident], chronic lung disease [including chronic obstructive pulmonary disease (COPD), asthma], chronic liver disease (including cirrhotic and non-cirrhotic), chronic kidney disease (CKD), and diabetes mellitus.

The recorded primary outcomes were hospitalization, intensive care unit (ICU) admission and 30-day all-cause mortality. Hospitalization was defined as within 2 days of a positive COVID test. All admissions included were to a medical, surgical or ICU treating specialty. Units and treating specialties coded specifically for rehabilitation or psychiatry were excluded. Deaths were identified as occurring within the VA facility or the VA was provided a death certificate, which was archived in the electronic Veteran registration package of VISTA. Extracted data also treating specialty and number of admissions.

The secondary outcome was post-vaccination infection (i.e. acquiring COVID-19 after being fully vaccinated), defined as first positive SARS-CoV-2 test two weeks after the second dose of the Pfizer-BioNTech or Moderna vaccine or one dose of the Janssen vaccine (starting on January 11, 2021).^26^ Vaccination status was defined “fully vaccinated” if Veterans had either received 2 doses of the Pfizer-BioNTech [BNT162b2 (Comirnaty)] or Moderna [mRNA-1273 (SpikeVAX)] vaccines, or one dose of the Janssen COVID-19 (Ad.26.COV2.S) vaccine. Vaccination status was defined based on documentation of vaccine receipt at one of the New England VHA sites or based on documented verification of vaccine receipt at a non-VA facility in the VA electronic health record by a VA staff member.

The majority of data originated from entry into the VistA electronic record system. This information is stored in the VA CDW, which requires use of SQL programming language. Several data subsets were created as tables, joined, and pivoted in order to meet meaningful use criteria. The application of SQL functions was used to accommodate the different reporting methods across sites, BMI, length of stay, most recent laboratory values, and various numerical results. Once all the data for this cohort was collated, it was analyzed and imported into statistical software.

### Sample handling and Genomic Sequencing

Handling of nasopharyngeal specimens and isolated virus was carried out by the VACHS clinical laboratory as part of clinical care, following standardized CLIA guidelines. Specimens were collected in viral transport medium, universal transport medium, or normal (0.9%) saline. RNA was extracted from fresh or frozen specimens using either the KingFisher Flex system (Thermo Fisher Scientific) or the QIAsymphony SP (QIAGEN), then shipped for whole genome sequencing (WGS) to a non-VA reference laboratory. In some cases, variant determinations were carried out on-site by multiplex PCR using the MassARRAY SARS-CoV-2 Variant Panel, Version 3 (Agena Bioscience).

We evaluated viral genomes from 951 Veterans: WGS was conducted on 723 viral genomes using Illumina (n = 224), Nanopore (n = 59), Illumina Nanoseq 6000 (n=358), and Illumina COVIDSeq (n = 82); while the MassARRAY system alone was used for variant determination on 228 Veteran specimens. Of the 951 total sequences, we had complete clinical data for 859 individuals. In cases where the MassARRAY gave indeterminate results or when the variant type was not included in the PCR panel, the specimens were reflexed to WGS. We correlated outcomes to identified variants, spike mutations, and lineages in 3 time periods: early (March – September 2020), middle (October 2020 – March 2021), and late (April 2021 – July 2021).

### Statistical Analysis

Logistic correlation analysis was used to find correlates of hospitalization, mortality, ICU admission, and post-vaccination infection. We stratified Veterans by vaccination status. We first conducted a univariate analysis, then used variables from the univariate analysis with *P* < 0.10 to use in a stepwise multivariate model with a forward selection level of 0.1 and backward selection level of 0.15. Z-tests were done to compare differences in proportions between vaccinated and unvaccinated hospitalizations. All analyses were performed using STATA v16 (College Station, TX).

## Results

### Veteran Characteristics

Of 4,170 Veterans in six New England states with confirmed SARS-CoV-2 during the study period, average age was 60.1 ±17.7 years, with 8% female and 81.1% White (Table 1a). An early cohort of 274 Veterans from our pilot study were included in this analysis to ensure the completeness of our analysis.^25^ The most common comorbidities were chronic heart disease (31%), diabetes (27%), and chronic lung disease (23%). Of the 2,399 Veterans who became fully vaccinated in our study, 53% received the Pfizer vaccine, 41% received Moderna, and 6% received Janssen (Table 1b).

**Table 1a.** Patient characteristics (n = 4,170)

| Demographics |  | Comorbidities |  |
| --- | --- | --- | --- |
| Age (years) | 60.1 ±17.7 | Dementia | 86 (12.06) |
| Sex |  | Alcohol use | 286 (6.86) |
| Male | 3,835 (92.0) | Tobacco use | 753 (18.1) |
| Female | 335 (8.03) | Drug use | 177 (4.24) |
| Race |  | Cannabis | 56 (1.34) |
| White | 3,320 (81.1) | Opioid | 25 (0.60) |
| Black | 526 (12.9) | Cocaine | 49 (1.18) |
| American Indian or Alaskan | 28 (0.68) | Other drugs | 102 (2.45) |
| Native Hawaiian/Pacific Islander | 15 (0.37) | Chronic Heart Disease | 1,289 (30.9) |
| Asian | 45 (1.10) | CAD | 633 (15.2) |
| Unknown/Declined | 159 (3.88) | Cardiomyopathy | 136 (3.26) |
| Ethnicity |  | Afib | 552 (13.2) |
| Non-Hispanic | 3,635 (88.8) | Other arrhythmia | 427 (10.2) |
| Hispanic | 272 (6.64) | CHF | 410 (9.83) |
| Unknown/Declined | 178 (4.61) | CVA | 137 (3.29) |
| State |  | Chronic Lung disease | 946 (22.7) |
| Massachusetts | 1,182 (43.7) | COPD | 701 (16.8) |
| Connecticut | 1,205 (28.9) | Asthma | 370 (8.87) |
| Maine | 258 (6.19) | Chronic liver disease | 350 (8.39) |
| New Hampshire | 377 (9.04) | Cirrhosis | 93 (2.23) |
| Rhode Island | 506 (12.1) | Non-cirrhotic | 312 (7.48) |
| Vermont | 2 (0.05) | CKD | 277 (6.64) |
| BMI category |  | Malignancy | 111 (2.66) |
| <25 | 467 (14.5) | Diabetes | 1,141 (27.4) |
| 25-30 | 1,041 (32.4) |  |  |
| 30-35 | 965 (30.0) |  |  |
| >35 | 739 (14.5) |  |  |

**Table 1b.**
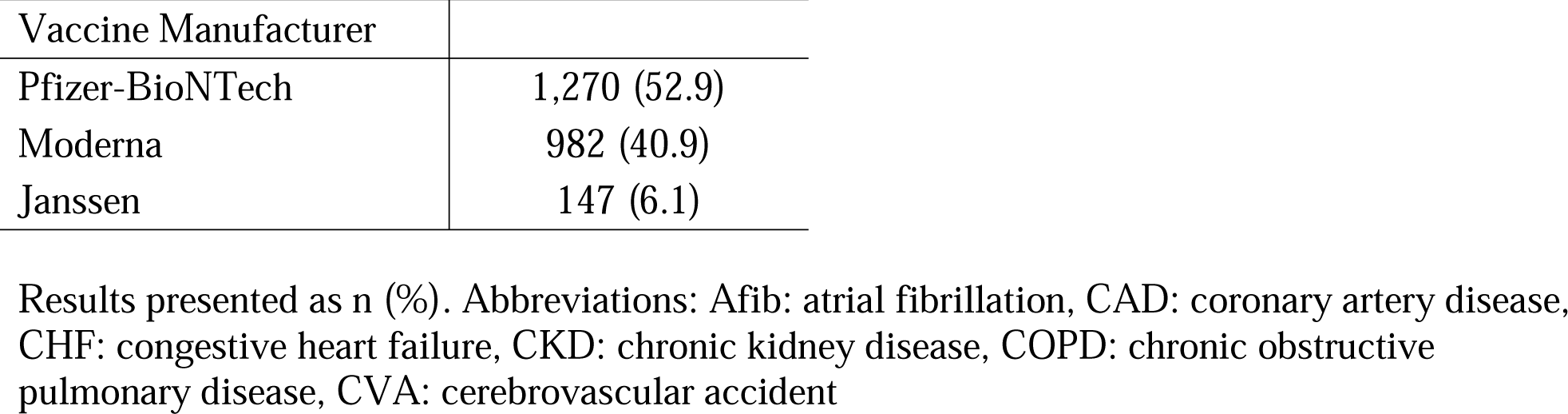
Vaccine Manufacturer among vaccinated patients (n = 2,399)

### Associations with outcomes before and after vaccination

Of the 4,170 Veterans who tested positive for COVID-19, a total of 811 Veterans were hospitalized (19%) (first hospitalization not fully vaccinated: n = 759, fully vaccinated: n = 52), with 118 having ICU admissions (2.8%) (not fully vaccinated: n = 112, fully vaccinated: n = 6), and 199 deaths (4.7%) (not fully vaccinated: n = 190, fully vaccinated: n = 9). Only 0.38% (9/2,399) of fully vaccinated patients died.

On multivariate regression (Table 2a), significant correlates of hospitalization in Veterans who first tested positive for SARS-CoV-2 before vaccination were age [odds ratio (OR): 1.05, 95% confidence interval (CI): 1.04, 1.06], male gender (OR: 1.63, 95% CI: 1.02, 2.61), black race (OR: 1.60, 95% CI: 1.28, 2.16), BMI of 30-34.9 (OR: 0.74,. 95% CI: 0.57, 0.97), chronic heart disease (OR: 1.64, 95% CI: 1.33, 2.01), COPD (OR: 1.39, 95% CI: 1.11, 1.75), chronic liver disease, 95% CI: OR: 1.43 (1.05, 1.94), and CKD (OR: 1.69, 95% CI: 1.24, 2.32). Significant correlates of ICU admission were age (OR: 1.04, 95% CI: 1.02, 1.05), BMI 30 – 34.9 (OR: 0.29, 95% CI: 0.15, 0.55), BMI 24 – 24.9 (OR: 0.48, 95% CI: 0.24, 0.94) and tobacco use (OR: 2.24, 95% CI: 1.41, 3.57). The only significant correlate of mortality was age (OR: 1.09, 95% CI: 1.07, 1.10).

**Table 2a.**
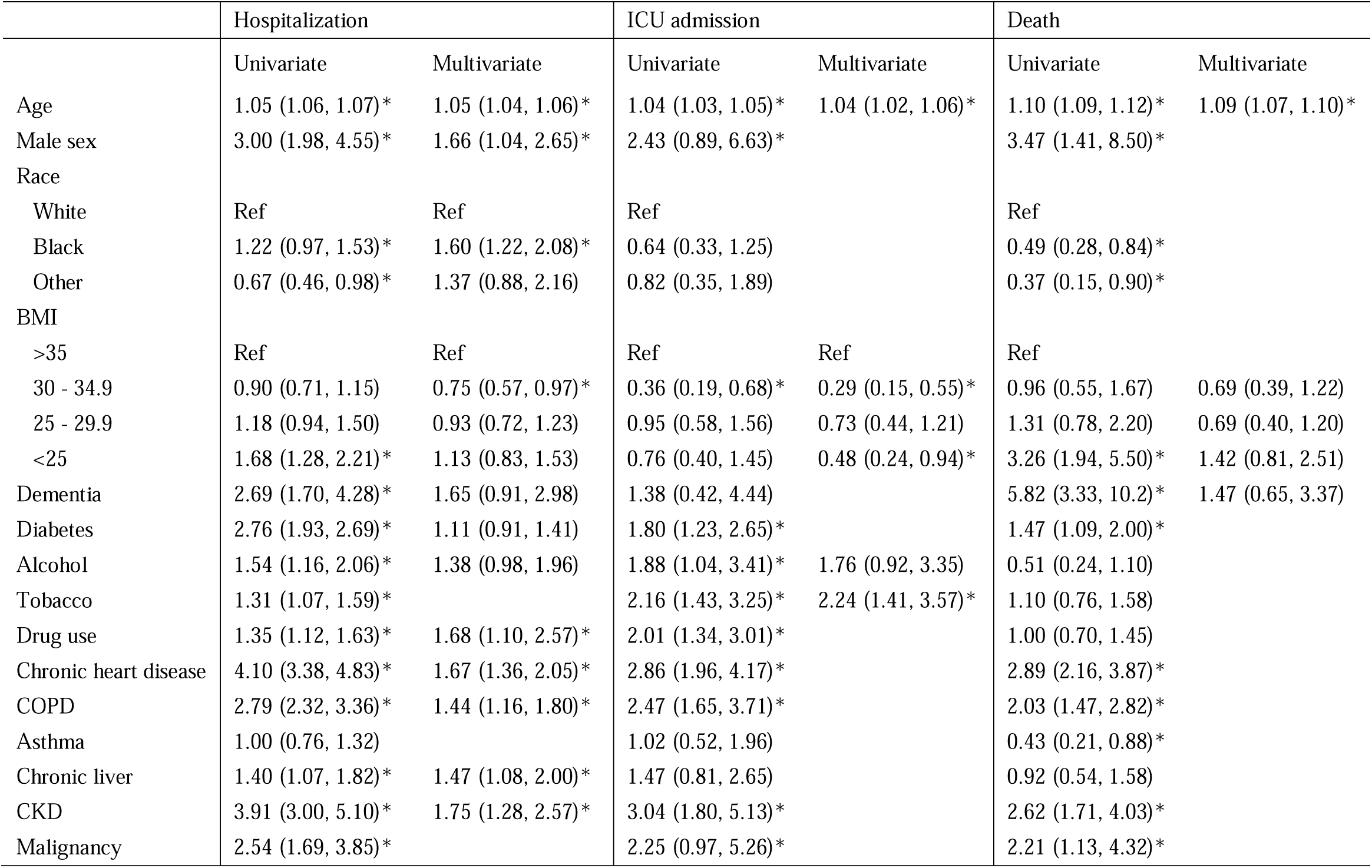
Regression analysis of correlates of hospitalization, ICU admission, and death in unvaccinated patients (n = 3,950)

**Table 2b.** Regression analysis of correlates of hospitalization in patients who first became COVID+ after full vaccination (n = 220)

|  | Hospitalization |  |
| --- | --- | --- |
|  | Univariate | Multivariate |
| Age | 1.03 (1.01, 1.05)* | 1.03 (1.00, 1.06)* |
| Male sex | 4.28 (0.55, 33.50) |  |
| Race |  |  |
| White | Ref |  |
| Black | 0.92 (0.32, 2.63) |  |
| Other | 0.89 (0.18, 4.46) |  |
| BMI |  |  |
| >35 | Ref |  |
| 30 - 34.9 | 0.78 (0.30, 2.00) |  |
| 25 - 29.9 | 0.77 (0.31, 1.90) |  |
| <25 | 1.56 (1.57, 4.31) |  |
| Dementia | 1.08 (0.211, 5.52) |  |
| Diabetes | 1.89 (1.01, 3.56)* |  |
| Alcohol | 2.30 (0.93, 5.68)* | 2.76 (1.06, 7.13)* |
| Tobacco | 0.86 (0.40, 1.83) |  |
| Drug use | 1.04 (0.51, 2.10) |  |
| Chronic heart disease | 2.11 (1.12, 3.96)* | 1.35 (0.67, 2.75) |
| COPD | 1.27 (0.61, 2.67) |  |
| Asthma | 0.94 (0.33, 2.69) |  |
| Chronic liver | 0.95 (0.33, 2.70) |  |
| CKD | 3.60 (1.65, 7.87)* | 3.26 (1.43, 7.47)* |
| Malignancy | 1.91 (0.54, 6.82) |  |

Results presented as odds ratio (95% confidence interval).
Abbreviations: BMI: body mass index, CKD: chronic kidney disease, COPD: chronic obstructive pulmonary disease

Out of Veterans who first tested positive for SARS-CoV-2 after full vaccination (n = 220) (Table 2b), significant correlates of hospitalization on multivariate analysis were age (OR: 1.03, 95% CI: 1.00, 1.06), alcohol use (OR: 2.76, 95% CI: 1.06, 7.13), and CKD (OR: 3.26, 95% CI: 1.43, 7.47).

### Relationship between vaccination and COVID hospitalization rates

A total of 220 (5.3% of total Veterans with positive COVID-19 test) Veterans tested positive for COVID-19 for the first time after being fully vaccinated, compared to the 3,950 Veterans (95%) who first tested positive when they were not vaccinated (difference of proportions: 0.90, 95% CI: 0.88, 0.92) (Figure 1). Of the 220 breakthrough cases, 24% (52 out of 220 cases) were hospitalized after testing positive the first time after vaccination, in comparison to the 19% of Veterans who tested positive for COVID-19 for the first time before vaccination and were hospitalized (759 out of 3,950 cases) (difference of proportions: 0.05, 95% CI: -0.10, 0.01).

**Figure 1.**
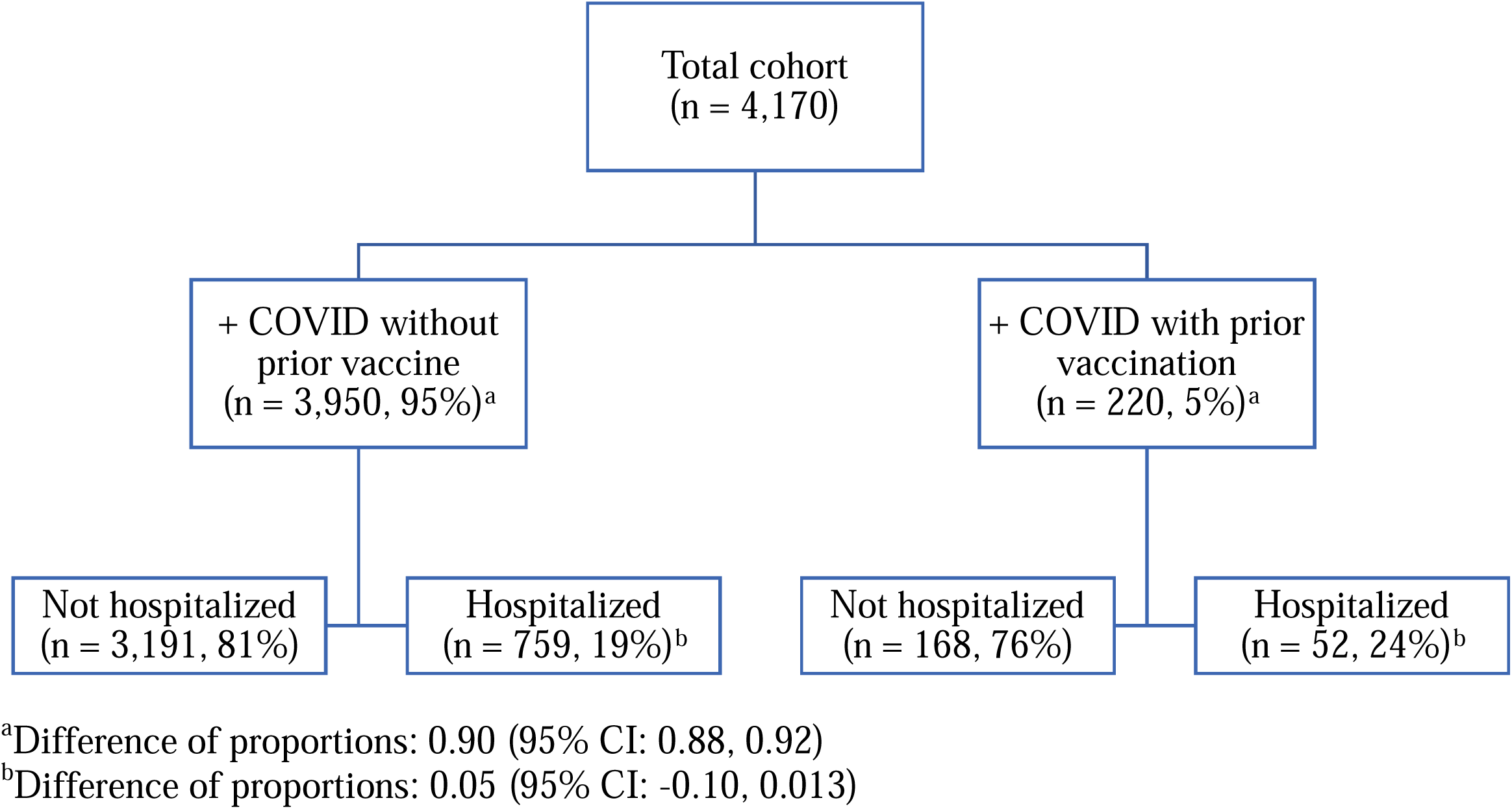
Hospitalization status by COVID vaccination status

There was a median of 111 days [25^th^ pctl: 52 days, 75^th^ pctl: 157 days) from being considered “fully vaccinated” (2 weeks after last dose) to the date of COVID-19 diagnosis. In regards to partially vaccinated Veterans, two hundred Veterans (4.8%) first tested positive for COVID after their first vaccination but before being fully vaccinated, with 53 Veterans hospitalized.

### Associations with first time Sars-CoV-2 positive test after full vaccination

Among the Veterans who first tested positive for COVID after being fully vaccinated (n = 220), the average age was 68.0 ± 15.3 years, 6.4% were female (n = 14), and 86% were white (n = 186), 127 received the Pfizer-BioNTech vaccine (58%, 10% of all Pfizer recipients), 83 received Moderna (38%, 8.5% of all Moderna recipients), and 10 received Janssen (4.5%, 6.8% of Janssen recipients) vaccine. Only 0.38% (9/2,399) of fully vaccinated Veterans died.

On multivariate analysis, we found that Veterans who tested positive for COVID-19 after being fully vaccinated (n = 220) (Supplemental Table 1), when compared to Veterans who were not fully vaccinated, were significantly more likely to be older (OR: 1.02, 95% CI: 1.02, 1.04), and had higher rates of tobacco use (OR: 1.50, 95% CI: 1.11, 2.03), drug use (OR: 1.79, 95% CI: 1.06, 3.02), CKD (OR: 1.59, 95% CI: 1.07, 2.35), and malignancy (OR: 1.78, 95% CI: 1.02, 3.11). No significant differences were found based on vaccine manufacturer.

### Viral Sequencing

From the WGS data, we found that New England Veterans (>97%) were infected with B.1 and its sub-lineages with the D614G mutation in the early (n = 282) and middle period (n = 349) of the study.^25^ In the late period (n = 277), the delta (B.1.617.2, 71%) lineages predominated (Supplemental Table 2). The most common and prominent lineages and subsequent hospitalizations within those lineages are shown in Figure 2. In the late period, Veterans who were infected after vaccination (n = 102) were more likely to have been infected with the delta variant (OR: 2.56, 95% CI: 1.41, 4.65) over other variants, but there was no difference in rates of being infected with the Delta variant over other variants based on vaccine status (p = 0.20). The most common spike mutations were D614G (n = 900), L452R (n = 245), T487K (n = 231), P231 (n = 231), and T19R (n = 230) (Supplemental Table 3).

**Figure 2.**
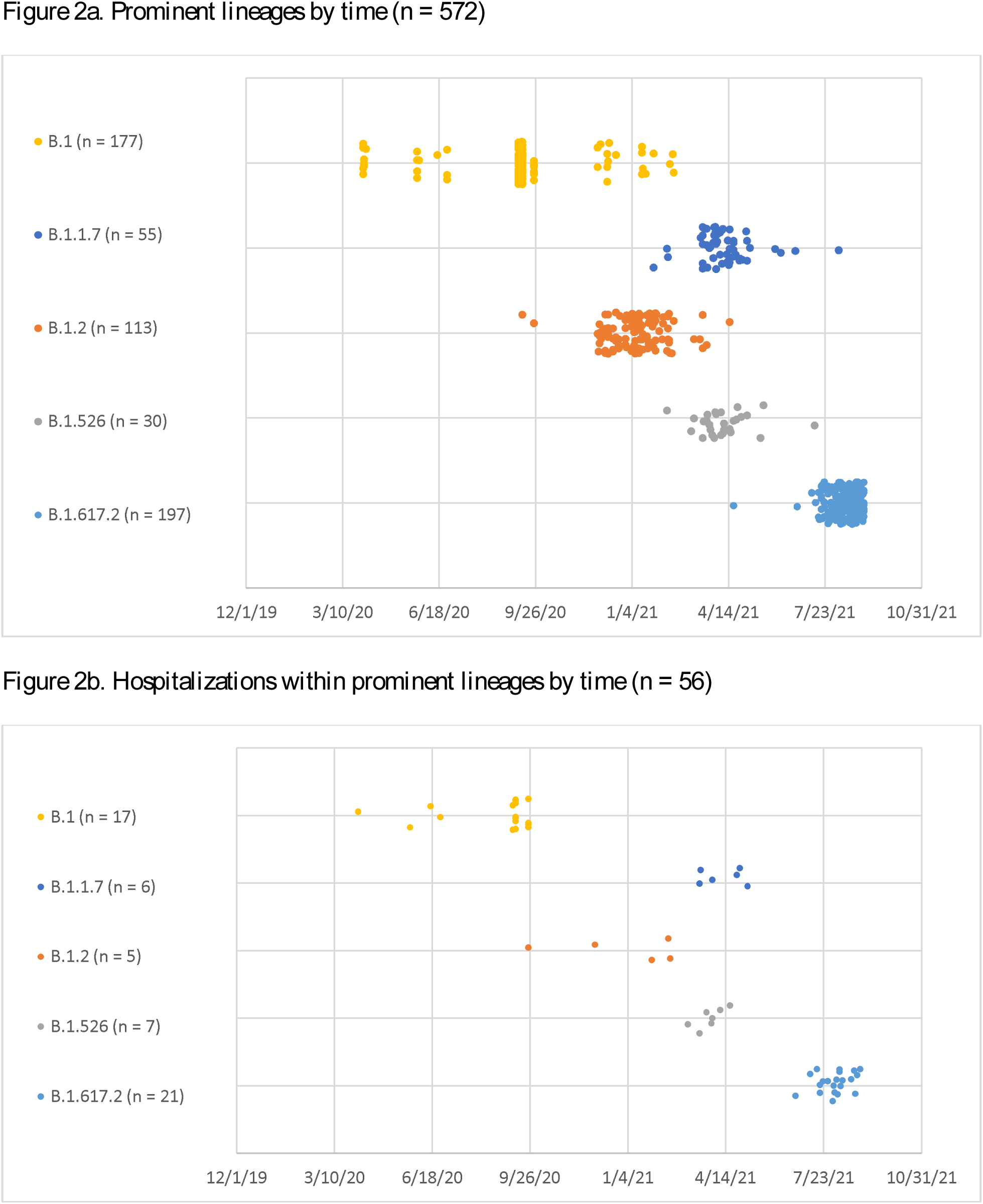
Graph of lineages and hospitalizations within lineages by time

## Discussion

Our study found that in an older cohort of Veterans with a high comorbidity burden, age significantly correlated with hospitalization, ICU admission, and mortality in unvaccinated populations. Drug use, tobacco use, CKD, and malignancy were higher in Veterans who contracted post-vaccination infection, irrespective of vaccination type. Among those who were infected post vaccination, age, alcohol use, and CKD correlated with hospitalization. The most common lineages in 2020 were B.1 and B.1.2, while B.1.617.2 dominated infections during the summer and fall of 2021. Our study is unique in that it includes comprehensive data from all Veterans in six New England states and additionally characterizes post-vaccination infection covariates. Veterans in our study are unique in that they are an older cohort with high rates of comorbid disease,^21,27^ and understanding clinical factors that impact outcomes in this population should help clinicians determine risks for Veterans with similar demographics.

Our data showed that starting in the summer of 2021, the B.1.617.2 (delta) variant dominated the New England Veteran infections. This lineage did not associate with worse hospitalization, ICU admission, or mortality in our study, but more people who were infected after vaccination in the late period were more likely to have this variant. Laboratory studies have shown the vaccine to be effective against the delta variant, although perhaps less so than other variants ^16,28^, and one clinical study from the United Kingdom noted modest differences in vaccine effectiveness from the delta variant as composed to the alpha variant.^29^ However, these latter findings must be taken into context that as delta becomes the predominant variant,^20,30^ vaccine-induced immunity (especially among patients vaccinated early in 2021) may be waning.^17^ Thus, ongoing efforts to better understand the duration of vaccine effectiveness and to define the need and timing for booster vaccinations are critical.^31^

Among fully vaccinated Veterans, 9% of fully vaccinated Veterans had breakthrough cases of COVID-19. This mirrors an increase in breakthrough cases throughout the globe.^18-20,32^ Our study showed a statistically significant higher risk of getting infected before vaccination, as compared to after vaccination, and the overall COVID-19 related hospitalizations were lower among the fully vaccinated. However, among those who did get COVID-19 after being fully vaccinated, the hospitalization rate was not statistically different compared to those who had it before vaccination. Given the data surrounding vaccine efficacy against COVID-19 hospitalization,^33,34^ this may indicate that the severity of disease may be higher with the newest variants of COVID-19 or that we did not have sufficient power to detect a difference. We did not detect a difference in breakthrough infection in our vaccine-recipient cohort rates by vaccine manufacturer, in contrast to other studies.^35^ Nevertheless, continued efforts to increase vaccination rates and encourage social distancing and mask use indoors are critical to curb infection spread.^18,19^

The comorbidities correlating with worse outcomes in our study were similar to analyses in some other populations of patients with COVID-19. Like other studies, we found that age was a significant correlate of worse outcomes.^4,7,36-39^ Male sex, which associated hospitalization in our study, is another widely cited predictor of worse prognosis,^22,24^ with one meta-analysis citing a global odds ratio of 2.84 for intensive treatment unit admission and 1.39 for higher odds of death.^24^ Our data also showed an increased rate of hospitalization among unvaccinated infected black participants, as has been shown in some previous studies, but this effect was lost in vaccinated black patients.^40,41^ In our univariate analysis, we found that Veterans with dementia had a higher risk of hospitalization and death, which has been replicated in other studies,^37,42,43^ but this effect was lost on multivariable analysis. Thus, the increased risk of dementia for poor clinical outcomes may be explained by age and comorbidities but nevertheless emphasizes the importance of additional monitoring required when caring for a patient with dementia; providers and caregivers must ensure this vulnerable population receives sufficient support throughout the course of their illness. Age, CKD, malignancy, drug use, and alcohol use were shown to be risk factors for getting infected among the vaccinated Veterans, with age, CKD and alcohol use being correlated most to hospitalization in this population. An understanding of clinical variables impacting risks among the vaccinated is critical to inform the vaccine-booster prioritization guidelines, and triaging guidelines for healthcare systems.

Limitations of this work include that it is specific to Veterans, a largely older male cohort, and results may not be generalizable to other populations. Evaluation of Veteran population is critical, however, given that these sex and age limitations also are known associations for COVID-19 infection. Our study spanned the course of the COVID-19 pandemic since its beginning in our region, and although triage algorithms in our hospital system did not significantly change over this time period, our study encompasses both pre- and post-vaccination time periods. We therefore analyzed outcomes in fully vaccinated Veterans and compared to those among unvaccinated. We were only able to record hospitalizations within the VA system and did not have results if Veterans were seen at outside hospitals. Furthermore, we are limited by the retrospective nature of our review, and analysis via ICD codes may have missed diagnoses due to incorrect or inaccurate coding. We did not have data on patients who did not test positive for COVID-19; future work would report COVID-19 positivity based on vaccine status/type. Post-vaccine hospitalizations had a shorter time to accumulate, and future studies should assess how time-based factors (including waves of the virus and likelihood of being hospitalized or infected over time) as well as vaccine uptake over time affect outcomes. Our study’s strengths include its comprehensive inclusion of all Veterans in New England, large study size, and novel analysis of correlates of post-vaccination infection.

## Conclusion

We found that in this regional cohort of Veterans with average age of 60 years and multiple comorbidities, age significantly correlated with hospitalization, ICU admission, and mortality. Veterans who tested positive for COVID-19 after vaccination were more likely to be older and had higher rates of tobacco use, drug use, CKD, and malignancy. This suggests that these populations may especially benefit from a booster dose. Our study also revealed that among fully vaccinated Veterans who become infected with COVID-19, age, alcohol use, and CKD associated with hospitalizations. This identifies vaccinated patients who test positive for COVID-19 most at risk for hospitalization and can inform triage algorithms. Hospitalization rates were similar among vaccinated and unvaccinated veterans, and mortality among vaccinated Veterans was rare. While the most common lineages in 2020 were B.1 and B.1.2, the delta variant of SARS-CoV-2 (B.1.617.2) dominated infections during the summer and early fall of 2021, and Veterans who were infected after vaccination were more likely to have this variant.

## Data Availability

All data produced in the manuscript are owned by the Veterans Affairs system.

## Acknowledgements

The authors thank Irina Tikhonova (Yale Center for Genome Analysis) and Nathan D. Grubaugh (Yale School of Public Health) for conduction and analysis of whole genome sequencing for some of the samples in the study. The authors also thank Danielle Plank (VA Connecticut Healthcare System) for extracting RNA for whole genome sequencing.

## Author Contributions

ML and SG participated in the conception, design, data collection, analysis and interpretation of results, and manuscript preparation. DR participated in the data collection, analysis and interpretation of results, and manuscript preparation. TCK participated in the analysis and interpretation of results and manuscript preparation. TC and GS participated in the conduction, analysis and interpretation of whole genome sequencing and in manuscript preparation.

