## Supplemental Table for "Clinical and Virologic Factors associated with Outcomes of COVID-19 before and after Vaccination among Veterans: Retrospective Analysis from Six New England States"

Supplemental Table 1. Regression analysis of patients who tested positive for COVID-19 after being fully vaccinated (n = 4,170)

|  | Univariate | Multivariate |
| --- | --- | --- |
| Age | 1.04 (1.03, 1.05)* | 1.02 (1.02, 1.04)* |
| Male sex | 3.58 (2.07, 6.18)* |  |
| Race |  |  |
| White | Ref |  |
| Black | 0.71 (0.45, 1.11) |  |
| Other | 0.52 (0.26, 1.02) |  |
| BMI |  |  |
| >35 | Ref |  |
| 30 - 34.9 | 0.91 (0.06, 1.40) |  |
| 25 - 29.9 | 1.00 (0.67, 1.51) |  |
| <25 | 1.06 (0.65, 1.75) |  |
| Dementia | 2.27 (1.08, 4.76)* |  |
| Diabetes | 2.17 (1.64, 2.87)* | 1.25 (0.96, 1.64) |
| Alcohol | 2.00 (1.28, 3.14)* |  |
| Tobacco | 1.74 (1.26, 2.41)* | 1.50 (1.11, 2.03)* |
| Drug use | 1.98 (1.18, 3.35)* | 1.79 (1.06, 3.02)* |
| Chronic heart disease | 1.32 (1.76, 3.05)* |  |
| COPD | 1.75 (1.26, 2.44)* |  |
| Asthma | 1.40 (0.89, 2.20) |  |
| Chronic liver | 1.50 (0.95, 2.37) |  |
| CKD | 3.15 (2.12, 4.68)* | 1.59 (1.07, 2.35)* |
| Malignancy | 2.46 (1.30, 4.66)* | 1.78 (1.02, 3.11)* |
| Vaccine kind |  |  |
| Pfizer-BioNTech | Ref |  |
| Moderna | 0.84 (0.62, 1.12) |  |
| Janssen | 0.63 (0.32, 1.23) |  |

*Results presented as odds ratio (95% confidence interval).*

Supplemental Table 2. Lineages by time period

| Early Period (n = 282)  (March 1 – Sept 30, 2020) | | | Middle Period (n = 349)  (Oct 1, 2020 – March 31, 2021) | | | Late Period (n = 277)  (April 1, 2021 – July 31, 2021) | | |
| --- | --- | --- | --- | --- | --- | --- | --- | --- |
| Lineage | Frequency | Percent | Lineage | Frequency | Percent | Lineage | Frequency | Percent |
| A | 1 | 0.35 | B.1 | 18 | 5.16 | P.1 | 1 | 0.36 |
| A.1 | 3 | 1.06 | B.1.1 | 4 | 1.15 | B.1.1.222 | 1 | 0.36 |
| A.2 | 2 | 0.71 | B.1.1.192 | 4 | 1.15 | B.1.1.7 | 34 | 12.27 |
| A.3 | 1 | 0.35 | B.1.1.222 | 2 | 0.57 | B.1.2 | 1 | 0.36 |
| B.1 | 159 | 56.38 | B.1.1.265 | 2 | 0.57 | B.1.319 | 1 | 0.36 |
| B.1.1 | 1 | 0.35 | B.1.1.28 | 1 | 0.29 | B.1.427 | 3 | 1.08 |
| B.1.1.10 | 1 | 0.35 | B.1.1.304 | 1 | 0.29 | B.1.429 | 1 | 0.36 |
| B.1.1.113 | 1 | 0.35 | B.1.1.316 | 13 | 3.72 | B.1.525 | 1 | 0.36 |
| B.1.1.128 | 1 | 0.35 | B.1.1.434 | 2 | 0.57 | B.1.526 | 10 | 3.61 |
| B.1.104 | 4 | 1.42 | B.1.1.464 | 1 | 0.29 | B.1.526 | 3 | 1.08 |
| B.1.108 | 1 | 0.35 | B.1.1.486 | 4 | 1.15 | B.1.526.1 | 2 | 0.72 |
| B.1.110.3 | 1 | 0.35 | B.1.1.7 | 21 | 6.02 | B.1.526.2 | 5 | 1.81 |
| B.1.117 | 1 | 0.35 | B.1.110.3 | 31 | 8.88 | B.1.617 | 1 | 0.36 |
| B.1.2 | 2 | 0.71 | B.1.139 | 3 | 0.86 | B.1.617.2 | 197 | 71.12 |
| B.1.268 | 1 | 0.35 | B.1.2 | 110 | 31.52 | B.1.621 | 2 | 0.72 |
| B.1.284 | 1 | 0.35 | B.1.234 | 1 | 0.29 | P.1 | 10 | 3.61 |
| B.1.302 | 26 | 9.22 | B.1.240 | 2 | 0.57 | P.1 | 1 | 0.36 |
| B.1.311 | 1 | 0.35 | B.1.243 | 6 | 1.72 | P.2 | 3 | 1.08 |
| B.1.320 | 1 | 0.35 | B.1.265 | 1 | 0.29 |  |  |  |
| B.1.356 | 18 | 6.38 | B.1.298 | 1 | 0.29 |  |  |  |
| B.1.359 | 7 | 2.48 | B.1.324 | 2 | 0.57 |  |  |  |
| B.1.36.10 | 1 | 0.35 | B.1.346 | 1 | 0.29 |  |  |  |
| B.1.36.8 | 1 | 0.35 | B.1.349 | 4 | 1.15 |  |  |  |
| B.1.369 | 3 | 1.06 | B.1.36.10 | 1 | 0.29 |  |  |  |
| B.1.382 | 1 | 0.35 | B.1.361 | 2 | 0.57 |  |  |  |
| B.1.385 | 1 | 0.35 | B.1.369 | 6 | 1.72 |  |  |  |
| B.1.390 | 16 | 5.67 | B.1.375 | 9 | 2.58 |  |  |  |
| B.1.403 | 1 | 0.35 | B.1.409 | 4 | 1.15 |  |  |  |
| B.1.413 | 1 | 0.35 | B.1.427 | 4 | 1.15 |  |  |  |
| B.1.417 | 1 | 0.35 | B.1.427 | 1 | 0.29 |  |  |  |
| B.1.422 | 1 | 0.35 | B.1.429 | 6 | 1.72 |  |  |  |
| B.1.436 | 1 | 0.35 | B.1.433 | 5 | 1.43 |  |  |  |
| B.1.443 | 1 | 0.35 | B.1.448 | 6 | 1.72 |  |  |  |
| B.1.479 | 10 | 3.55 | B.1.478 | 1 | 0.29 |  |  |  |
| B.1.509 | 1 | 0.35 | B.1.509 | 1 | 0.29 |  |  |  |
| B.1.517 | 3 | 1.06 | B.1.517 | 33 | 9.46 |  |  |  |
| B.1.521 | 1 | 0.35 | B.1.526 | 9 | 2.58 |  |  |  |
| B.1.590 | 1 | 0.35 | B.1.526 | 1 | 0.29 |  |  |  |
| B.1.595.3 | 1 | 0.35 | B.1.526.2 | 4 | 1.15 |  |  |  |
| B.35 | 1 | 0.35 | B.1.541 | 1 | 0.29 |  |  |  |
| N.1 | 1 | 0.35 | B.1.575 | 1 | 0.29 |  |  |  |
|  |  |  | B.1.577 | 1 | 0.29 |  |  |  |
|  |  |  | B.1.595 | 2 | 0.57 |  |  |  |
|  |  |  | B.1.596 | 3 | 0.86 |  |  |  |
|  |  |  | B.1.603 | 3 | 0.86 |  |  |  |
|  |  |  | B.1.609 | 2 | 0.57 |  |  |  |
|  |  |  | None | 1 | 0.29 |  |  |  |
|  |  |  | P.1 | 2 | 0.57 |  |  |  |
|  |  |  | P.2 | 1 | 0.29 |  |  |  |
|  |  |  | R.1 | 3 | 0.86 |  |  |  |
|  |  |  | R.2 | 1 | 0.29 |  |  |  |

Supplemental Table 3. Most common Spike mutations

| Mutation | Frequency | Percentage |
| --- | --- | --- |
| D614G | 858 | 94.5 |
| L452R | 208 | 22.9 |
| T478K | 197 | 21.7 |
| P681R | 197 | 21.7 |
| T19R | 196 | 21.6 |
| N501all | 92 | 10.1 |
| Q677H | 69 | 7.6 |
| T95I | 61 | 6.7 |
| N501Y | 57 | 6.3 |
| P681H | 53 | 5.8 |
